# Effectiveness, barriers, and facilitating factors of strategies for active delabeling of patients with penicillin allergy labels: a systematic review protocol

**DOI:** 10.1101/2023.07.17.23291645

**Authors:** Hannah Nürnberg, Elham Khatamzas, Claudia M. Denkinger, Tabea Krause, Lars Oetken, Sophie Rauer, Amelie Rapp, Torsten Hoppe-Tichy, Benedict Morath

**Affiliations:** Hospital Pharmacy, University Hospital Heidelberg, Heidelberg, Germany; Division of Infectious Diseases and Tropical Medicine, University Hospital Heidelberg, Heidelberg, Germany; German Center of Infection Research, Partner site Heidelberg, Germany

**Keywords:** penicillin allergy delabeling, antibiotic stewardship

## Abstract

**Introduction:** Up to 15 % of adult patients in the clinical setting report to be allergic to penicillin. However, in most cases penicillin allergy is not confirmed. Due to the negative aspects associated with erroneous penicillin allergy, the implementation of active delabeling processes for penicillin allergy is an important part of antibiotic stewardship programs. Depending on the clinical setting, different factors need to be considered during implementation. This review examines the effectiveness of different delabeling interventions and summarizes components and structures that facilitate, support, or constrain structured penicillin allergy delabeling.

**Methods and analysis:** This review will adhere to the Preferred Reporting Items for Systematic Reviews and Meta-Analyses (PRISMA). The Databases MEDLINE (via PubMed), EMBASE, and Cochrane Library were searched for studies reporting on any intervention to identify, assess, or rule out erroneous penicillin allergy. Study design, intervention type, professional groups involved, effectiveness, limitations, barriers, facilitating factors, clinical setting, and associated regulatory factors will be extracted and analyzed. Two independent reviewers will perform the screening process and data extraction. Discordant decisions will be resolved through review by a third reviewer. Bias assessment of the individual studies will be performed using the Newcastle Ottawa Scale.

**Ethics and dissemination:** Because individual patient-related data is not analyzed, an ethical approval is not required. The review will be published in a peer-reviewed scientific journal.

**STRENGHTS AND LIMITATIONS OF THIS STUDY:**

- The systematic review will adhere to the PRISMA guidelines.
- A wide search strategy is used and the search will be conducted on three major databases.
- The search is focused on studies reporting on facilitators and barriers for implementation as well as effectiveness of penicillin allergy delabeling interventions.
- Data extraction will be performed using an established second look process.
- Risk for bias in the individual studies as well as external validity is assessed using an established checklist (Newcastle Ottawa Scale).

## INTRODUCTION

### Rationale

Penicillin allergy is the most commonly documented drug allergy in medical records [1]. In developed countries 5 % to 15 % of patients report to be allergic to penicillin [2]. However, less than 10 % of these patients will have a proven penicillin allergy [3]. Consequently, a penicillin allergy label can lead to life-long avoidance of beta-lactam antibiotics with increased prescription rates of second line antibiotics in this patient population [4, 5]. Compared with penicillin antibiotics, these antibiotics are associated with higher rates of adverse drug reactions, increased length of hospital stay, higher treatment costs, and in addition promote the emergence of antibiotic resistance [6-10]. Further, second line antibiotics often show lower efficacy, increasing the risk of treatment failure as well as mortality [11]. Large studies have shown that patients with documented penicillin allergy have a higher prevalence of *Clostridium difficile*, methicillin-resistant *Staphylococcus* aureus (MRSA), or vancomycin-resistant Enterococci (VRE) [12, 13]. In addition, the psychological impact due to allergies to antibiotics such as penicillin on patients is thought to be significant [14].

Because of these adverse aspects, structured testing of the diagnosis “penicillin allergy” prior to the prescription of an alternative antibiotic is an important tool in antibiotic stewardship programs to avoid the unnecessary use of second line antibiotics.

In order to provide penicillin allergy delabeling interventions in daily clinical routine, thorough and individualized process implementation appears to be of great importance [15]. Different clinical settings have individual structures that need to be considered during the implementation of complex interventions [15]. In addition, settings are specific to different countries mainly due to regulatory differences, e.g. pharmacists prescribing is well established in some but not all settings [16-18]. Different competences of professional groups involved in the intervention may therefore have implications for the applicability of penicillin allergy delabeling interventions and should be considered during implementation.

The goal of this review is to search relevant studies for effectiveness of interventions for penicillin allergy delabeling, limitations, barriers, and facilitating factors specific to the studied clinical setting and analyze them with the goal to create a toolbox of interventions that supports the implementation of penicillin allergy delabeling process most suited to different settings.

### Objectives

Previously published systematic reviews primarily focused on the safety of various delabeling methods, their feasibility by different professional groups, or perform a cost analysis [9, 19, 20]. The purpose of this systematic review is to expand the perspective on implementation and effectiveness, and to summarize components and structures that enable, promote, or constrain structured penicillin allergy delabeling. Based on this approach, applicable concepts to assist in the development and implementation of interprofessional strategies for active penicillin allergy delabeling will be provided.

### Review question

How are penicillin delabeling processes implemented in clinical practice and what are the reported effectiveness, barriers, and facilitating factors of implementation?

## METHODS AND ANALYSIS

### Overview

This systematic review protocol adheres to the Preferred Reporting Items for Systematic Reviews and Meta-Analyses Protocols (PRISMA-P) guideline and the systematic review will follow to the Preferred Reporting Items for Systematic Reviews and Meta-Analyses (PRISMA) guideline [21, 22]. This systematic review examines structural components and processes applicable for active delabeling of patients with documented penicillin allergy. Further, this review analyzes factors and determinants that are reported to support or limit the implementation of such processes in clinical routine.

### Inclusion criteria

For this systematic review, prospective studies are eligible that include adult patients in any clinical setting with a documented penicillin allergy.

The following prospective study designs will be eligible for inclusion: Randomized and non-randomized trials, before and after studies, prospective cohort studies, case-control studies, ecological studies, and quality improvement studies. Only studies available in English or German will be included.

Eligible studies need to report on any intervention to identify, assess, or rule out erroneous penicillin allergy. Due to the study designs and the mode of intervention, it is expected that there will be studies without a comparison group. Therefore, a comparison group is neither specified in the inclusion nor in the exclusion criteria to not limit results. Studies that analyzed the following outcomes are determined eligible. The primary outcome is the number of patients, who were successfully cleared from their penicillin allergy label due to the delabeling intervention or comparable measures evaluating the overall effect of the intervention (e.g. odds ratio). To expand the perspective on implementation and process, the following secondary outcomes are eligible for inclusion: (1) Barriers and facilitating factors for the implementation of penicillin allergy delabeling interventions as well as (2) measurable impact on healthcare summarized by potential markers of healthcare utilization such as incidence of postoperative infections associated with penicillin allergy, length of stay, treatment costs, changes in antibiotic use, and doses of broad spectrum antibiotics.

### Exclusion criteria

Studies that include participants younger than 18 years or not hospitalized patients will be excluded as well as studies mainly conducted in primary care. Case reports, letters, editorials, and reviews, but also studies that report on penicillin allergy testing of patients without a penicillin allergy label (in context of the primary diagnosis) are not eligible. However, all reviews identified during the search will be screened for eligible studies. There are no defined outcomes that will lead to an exclusion.

### Information sources

The databases PubMed/Medline, EMBASE and Cochrane Library will be searched from 1992 to 2023 for peer-reviewed scientific literature.

### Search strategy

The search strategies used for the different databases is provided in Table 1. To finalize the search strategy, various keywords and Medical Subject Headings (MeSH terms)/Emtree terms were first identified from relevant literature and explored in different databases. Subsequently, the combined search terms were defined by a consensus-based process (four-eyes principle). The key search concepts “penicillin allergy” and “delabeling” are consistently used as main keywords and terms. The other terms are translated and adapted to the available terms of the respective database.

**Table 1.** Search strategy in the databases.

| PubMed/Medline (searched on 06 June 2023) |  | Items found |
| --- | --- | --- |
| #1 | ("Penicillins/adverse effects"[Mesh] OR "Penicillins/drug effects"[Mesh])<br>OR ("beta-Lactams/adverse effects"[Mesh] OR "beta-Lactams/drug<br>effects"[Mesh]) | 15 252 |
| #2 | ("penicillin"[tw] OR "beta-lactam"[tw] OR "beta lactam"[tw]) AND<br>("allerg*" [tw] OR "reaction"[tw]) | 9 811 |
| #3 | ( "Drug Hypersensitivity/diagnosis"[Mesh] OR "Drug<br>Hypersensitivity/drug therapy"[Mesh] OR "Drug<br>Hypersensitivity/prevention and control"[Mesh] OR "Drug<br>Hypersensitivity/therapy"[Mesh] OR "Drug Hypersensitivity/organization<br>and administration"[Mesh]) OR "Delabeling"[tw] OR "Delabelling"[tw] OR<br>"oral challenge*" [tw] OR "Allergy test*" [tw] OR "management"[tw] OR<br>"intervention*" [tw] OR "penicillin testing"[tw] OR "penicillin test*" [tw]<br>OR "provocation test*" [tw] OR "provocation"[tw] OR "amoxicillin<br>challenge"[tw] OR "penicillin challenge"[tw] | 2 802 624 |
| #4 | #1 OR #2 | 22 722 |
| #5 | #3 AND #4<br>Publication Date: 1992-2023 | 2419 |
| Cochrane Library (searched on 06 June 2023) |  | Items found |
| #1 | "Penicillins" [MeSH] OR "beta lactam" [MeSH] | 10504 |
| #2 | "Drug Hypersensitivity" [MeSH] | 1380 |
| #3 | #1 AND #2 | 93 |
| #4 | Search terms: (Pencillin OR beta lactam OR beta-lactam) AND (allerg* OR reaction*) | 909 |
| #5 | #3 OR #4 | 967 |
| #6 | Search terms: Delabel*ing OR penicillin challenge OR amoxicillin challenge OR penicillin test* OR intervention* OR management OR provocation test* OR provocation OR oral challenge OR label*ing | 660 125 |
| #7 | #5 AND #6<br>Publication Date: 1992-2023 | 591 |
| EMBASE (searched on 06 June 2023) |  | Items found |
| #1 | 'beta lactam'/exp OR 'beta lactam' OR 'penicillin' OR 'penicillin derivative' | 209 858 |
| #2 | 'allergy' OR 'penicillin allergy' OR 'drug hypersensitivity' OR 'allergic reaction' | 505 757 |
| #3 | #1 AND #2 | 14 165 |
| #4 | 'delabeling' OR 'delabel*ing' OR 'amoxicillin challenge' OR 'penicillin challenge' OR 'penicillin test*' OR 'intervention' OR 'medication therapy management' OR 'provocation test' OR 'provocation test*' or 'oral challenge' | 1 316 139 |
| #5 | #3 AND #4<br>Publication Date 1992-2023 | 2007 |

### Study records

All identified publications will be collected using MS Excel® (Microsoft, Richmond) and duplicates resulting from searches on different databases will be removed prior the screening process. Three independent reviewers will review titles and subsequently abstracts of the search results according to the inclusion criteria. For inclusion, consensus between two reviewers is needed. In case consensus is not reached, an adjudicator will decide.

In a next step, the full text will be screened by two reviewers. Analogue to title and abstract screening, occurring disagreements during the screening process will be resolved by consensus or, if necessary, by a third reviewer. The process of study selection and the results of each screening step will be presented in a flow chart according to PRISMA.

### Data collection process

Data will be extracted to a predefined and standardized Excel sheet by one reviewer and reviewed by another (four-eyes principle). Any occurring discrepancies during the data collection process will be resolved by consensus or by a third reviewer. If necessary, the authors of the papers will be contacted to enquire missing or additional data.

Data will include information on the study design and the particular inpatient setting. The type of intervention (e.g. risk stratification, oral challenge, skin testing or combinations) will be extracted. In addition, the professional groups involved in the intervention will be collected and evaluated with regard to mentioned influence on the feasibility of the intervention. In terms of outcomes, effectiveness (measured by the number of successfully delabeled patients or comparable measures), reported limitations, barriers, and facilitating factors will be extracted. To evaluate transferability, associated regulatory factors (e.g. pharmacist prescribers available) will be analyzed.

### Risk of bias in individual studies

To assess the reliability of eligible studies, a bias assessment will be performed using the Newcastle Ottwawa scale [23]. The summary of findings will report on the proportion of patients successfully cleared from their penicillin allergy label, on barriers and facilitating factors for implementation of penicillin allergy delabeling intervention, and on any measured impact on healthcare.

### Data synthesis

No quantitative synthesis will be performed as this review aims on comprehensively characterizing available penicillin allergy delabeling concepts with a focus on intervention tools, structural components, barriers, and facilitators of implementation. Consequently, clinical settings, interventions, and professional groups will be described and characterized in terms to effectiveness of the delabeling process.

### Study status

On June 6^th^ 2023, literature was searched as previously described. Prior to final publication of the review, the search will be repeated to cover new studies published after June 06^th^. Currently, the studies are screened for inclusion and publication of the review is planned for the beginning of 2024.

### Strengths

This systematic review has several strengths. The systematic review will adhere to the PRISMA guidelines. To assess the certainty of the results, a thorough bias assessment of included studies will be performed by established tools targeting the study type. A wide search strategy is used including both text words and MeSH terms/Emtree terms. The search will be conducted on three major databases, increasing the likelihood that all relevant studies will be found. The systematic review is aimed to present a holistic view from effect to implementation of penicillin allergy delabeling interventions.

### Limitations

The systematic review has some limitations. Literature that can only be found in other databases, like Web of Science, might not be discovered. In addition, no sources were searched that contained unpublished or grey literature. Therefore, information from, for example, unpublished clinical trials, healthcare system reports, conference proceedings, or dissertations may not have been included. Furthermore, there may be relevant literature that is not available in English or German and therefore is not considered. In addition, no meta-analysis is performed and it appears likely that a majority of studies will be quasi-experimental studies, cohort studies, and single center studies that often are at a higher risk of bias. However, as this is the current available evidence, it is the aim to thoroughly collect the available data and provide an overview of the current penicillin delabeling landscape including aspects of implementation. Further, this risk and potential methodological flaws of included studies will be accounted for by a strict bias assessment.

## Data Availability

All data produced in the planned review will be available on reasonable request. Data sharing for the protocol not applicable

## ETHICS AND DISSEMINATION

Because this review does not analyze individual patient-related data and only secondary data, no ethical approval is required. It is planned to publish our systematic review in a peer-reviewed scientific journal.

## AUTHORS’ CONTRIBUTIONS

The protocol for the systematic review was developed by HN and critically edited and reviewed by BM, EK, THT and CMD. TK, LO, SR, and AR participated in the literature search and contributed to the methodological screening of the systematic review.

## PATIENT AND PUBLIC INVOLVEMENT STATEMENT

Patients or the public were not involved in the design, the conduct, or reporting, or dissemination plans of this research.

